# The Association of Fear of Movement and Postural Sway in People with Low Back Pain

**DOI:** 10.1101/2022.07.27.22278125

**Authors:** Anita Meinke, Cinzia Maschio, Michael Meier, Walter Karlen, Jaap Swanenburg

## Abstract

**Background:** Fear of movement is thought to interfere with the recovery from low back pain (LBP). To date, the relationship between fear of movement and postural balance has not been adequately elucidated. Recent findings suggest that more specific fears need to be assessed and put in relation to a specific movement task. We propose that the fear to bend the trunk in a certain direction is distinctly related to the amount of postural sway in different directions. Therefore, our aim was to investigate whether fear of movement in general and fear of bending the trunk in a certain plane is related to postural sway.

**Methods:** Data was collected from participants with LBP during two assessments approximately three weeks apart. Postural sway was measured with a force-platform during quiet standing with the eyes closed. Fear of movement was assessed with an abbreviated version of the Tampa Scale of Kinesiophobia (TSK-11) and custom items referring to fear of bending the trunk in the sagittal and the frontal plane.

**Results:** Based on data from 25 participants, fear of bending the trunk in the frontal plane was positively related to displacement in the sagittal and frontal plane and to velocity in the frontal plane (X^2^ =4.35, P=.04; X^2^ =8.15, P=.004; X^2^ =9.79, P=.002). Fear of bending the trunk in the sagittal plane was not associated with any direction specific measure of sway. A positive relation of the TSK-11 with velocity of the frontal plane (X^2^ =7.14, P=.008) was found, but no association with undirected measures of sway.

**Discussion:** Fear of bending the trunk in the frontal plane may be especially relevant to postural sway under the investigated stance conditions. It is possible that fear of bending the trunk in the frontal plane could interfere with balance control at the hip, shifting the weight from side to side to control balance.

**Conclusion:** For the first time the directional relationship of fear of movement and postural sway was studied. Fear of bending the trunk in the frontal plane was positively associated with several measures of postural sway.

## 1 Introduction

The estimates of “disability” for low back pain (LBP) generated within the global burden of disease study are larger than for any other complaint (Wu et al., 2020). Furthermore, a relationship between disability questionnaires and pain-related fear in people with LBP (Costa et al., 2011; Carvalho et al., 2017; Nordstoga et al., 2019) or other similar pain conditions (Luque-Suarez et al., 2019) is often observed. These connections highlight the need to understand the involvement and consequences of pain-related fear for people with LBP.

For many years, it was suspected that pain-related fears lead to adverse outcomes in people with LBP (Vlaeyen et al., 1995, Vlaeyen and Crombez, 1999). Pain-related fears are a cluster of multiple related concepts (Leeuw et al., 2007). This includes fear of movement, which can be due to the expectation that a movement may cause harm to the back, rather than fear of the pain itself (Vlaeyen et al., 1995). The basic premise of the work centered around fear of movement is that some people respond to pain with fear and withdraw from activities they regard as potentially harmful (Vlaeyen et al., 1995, Vlaeyen and Crombez, 1999). Consequently, they could lose the benefits of physical activity and movement, and might consolidate their pain even further (Vlaeyen et al., 1995, Vlaeyen and Crombez, 1999).

Indeed, a review showed that stronger fear of movement and avoidance beliefs during subacute LBP may promote difficulties to return to the office (Wertli et al., 2014). However, not all data available are in line with this model (Pincus et al., 2010; Costa et al., 2011). For example, Carvalho et al. (2017) did not observe a reduction in physical activity with increased fear of movement. On the other hand, LBP is linked to movement behavior and while it is assumed that these altered movements may serve to avoid physical stress on the spine or surrounding tissues, they could possibly cause additional physical stress (van Dieën et al., 2019). For example, a protective movement behavior (i.e., stiffening the spine) might be associated with increased muscle co-contraction, potentially causing additional load on spinal structures (van Dieën et al., 2019). This additional physical stress may jeopardize back health (van Dieën et al., 2019). Van Dieën et al. (2019) also remarked that such reactions could be purely detrimental without any advantage for the person if they are provoked by unjustified fears. Thus, more detailed insight in the associations between fear of movement and actual movement behavior may become useful in the prevention and management of LBP.

An association between fear of movement and the tendency to move the trunk in a stiff manner is discussed in the scientific literature (Karayannis et al., 2013; Alsubaie et al., 2021; Christe et al., 2021). Recently, a meta-analysis found that people with LBP and greater fear of movement may only slightly restrain their movement of the spine (Christe et al., 2021). Karayannis et al. (2013) found higher rigidity in reaction to perturbations of the trunk, and flexion in the sagittal plane occurs more slowly when fear of movement is elevated (Nordstoga et al., 2019; Osumi et al., 2019). Furthermore, people with greater fear avoidance beliefs and LBP may be more imprecise in tracing a requested movement trajectory by flexing and extending their trunk (Alsubaie et al., 2021).

Assessments of postural balance are commonly used in studies investigating LBP. “Balance is a generic term describing the dynamics of body posture to prevent falling” (Winter, 1995, 194). The operation of the sensorimotor system regulating balance can be observed by describing the pathway of the vertical ground reaction force, also known as center of pressure (COP) (Winter, 1995). In other terms, the COP based variables describe the motor activity generated to steer the body’s sway and not the actual movement of the center of mass (Winter, 1995). Nevertheless, for consistency with other publications, we will discuss COP-based variables, as a proxy measure of body sway. The regulation of body sway relies on sensory information such as proprioceptive, visual, and vestibular signals (Schmidt et al., 2019). For example, if vision is removed, sway variables such as velocity or range may slightly increase (Roman-Liu, 2018). During quiet standing with approximately parallel feet, sway on the sagittal plane predominantly means rotation around the ankle and muscle activity at the ankle (Winter, 1995). Rotation at the hip also occurs but plays a smaller role under these conditions (Creath et al., 2005). On the frontal plane weight is shifted from one leg to the other and the hip musculature plays a larger role than the musculature at the lower legs (Winter, 1995).

Several reviews have summarized research results on sway in people with LBP (Ruhe et al., 2011; Mazaheri et al., 2013; Berenshteyn et al., 2019; Koch and Hänsel, 2019). Three of these reviews highlighted the enlarged sway variables, for example in for velocity or sway area found in people with LBP (Ruhe et al., 2011; Berenshteyn et al., 2019). Specifically, this was the case, when contributions from the visual sense were removed (Koch and Hänsel, 2019; Ruhe et al., 2011; Berenshteyn et al., 2019). In contrast, another review emphasized that a subset of studies found a reduction in sway in people with LBP (Mazaheri et al., 2013). This heterogeneity in study results could arise from counteracting mechanisms of pain-related fear and other mechanisms associated with pain, which are thought to increase sway (Mazaheri et al., 2013). Kiers et al. (2015) proposed that fear of pain opposes the effect of pain by increasing stiffness. All these aspects illustrate the need for further investigations of the impact of pain-related fear on sway assessments (Mazaheri et al., 2013; Berenshteyn et al., 2019).

There are already some studies that reported data on the association of pain-related fear assessments with postural sway or other balance measures in people with LBP (Mazaheri et al., 2014; Sung et al., 2015; Jacobs et al., 2016; Kahraman et al., 2018; Shanbehzadeh et al., 2018; Hlaing et al., 2020; Zhang et al., 2020; Mikkonen et al., 2022). Several of these studies investigated bipedal, quiet stance conditions (Mazaheri et al., 2014; Kahraman et al., 2018; Shanbehzadeh et al., 2018; Zhang et al., 2020; Mikkonen et al., 2022). Mazaheri et al. (2014) evaluated the assumptions of counteracting mechanisms and inferred from their findings that fear of movement did not impact sway. However, the association of fear of movement and sway was not directly analyzed (Mazaheri et al., 2014). Instead, the inference was made by comparison of a group of people who had just overcome LBP (as fear was not diminished in this group yet), to people with ongoing and without LBP (Mazaheri et al., 2014). Kahraman et al. (2018) found a positive association with a measure combining sway across different manipulations of sensory input in male, but not in female participants or for any of the individual conditions. Similarly, Mikkonen et al. (2022) presented results from a larger group of participants, who were assessed under different conditions in narrow stance and several small but significant positive correlations of fear of movement and postural sway area were reported. However, these associations were not found for postural sway velocity, for which age alone was a significant predictor in a model including additional variables (Mikkonen et al., 2022). The authors concluded that there was no association between fear of movement and postural sway (Mikkonen et al., 2022). In contrast with the studies mentioned before, Shanbehzadeh et al. (2018) found that participants with greater pain-related fear showed decreased postural sway across multiple conditions including manipulations of vision, proprioception, and cognition. Unlike the other studies (Mazaheri et al., 2014; Kahraman et al., 2018; Mikkonen et al., 2022), which had used the Tampa scale of Kinesiophobia (TSK), Shanbehzadeh et al. (2018) used the Pain Anxiety Symptoms Scale. In addition to fear of movement, catastrophic thoughts in people with chronic LBP appear to be also linked to sway, although the direction of this association was reported inconsistently (Zhang et al., 2020).

Other researchers have used different balance assessments, such as standing on a single leg (Kahraman et al., 2018; Hlaing et al., 2020), sitting (Sung et al., 2015) or assessments with surface perturbation (Jacobs et al., 2016). In the study of Kahraman et al. (2018), fear of movement was negatively associated with some limits of stability measures, but they found no association with fear of movement for sway measures in single legged stance. Hlaing et al. (2020) neither found a relation of fear of movement with the time participants could maintain single leg stance on firm ground but observed a negative association on compliant ground. In a cohort of people who had not yet developed chronic LBP and who displayed reduced motor control capabilities during movement assessments, balance during sitting on a platform which was only supported in the center, was not found to correlate with fear avoidance beliefs (Sung et al., 2015). In contrast, another study that investigated how much the body shifted when reacting to tilts of the supporting ground, showed a positive relation with fear avoidance beliefs (Jacobs et al., 2016). Thus, there is some first evidence for a relation between fear of movement and postural sway, although several studies found no convincing association. Nevertheless, the current evidence appears difficult to reconcile. The differing results could be due to methodological variations, such as differences between balance and pain-related fear assessments.

It has been argued that commonly used comprehensive assessments of fear of movement do not capture fear to perform distinct movements (Leeuw et al., 2007) and thus make it harder to detect associations between fear of movement and the investigated movement (Pincus et al., 2010; Matheve et al., 2019). Indeed, Matheve et al. (2019) only identified a negative association with movement in the lumbar region during lifting with a measure directly quantifying fear of lifting, but not with common assessment tool for fear of movement. Further research confirmed analogous findings in people without pain (Knechtle et al., 2021) and by referring to such results, other researchers emphasized that more targeted assessments should be used (Christe et al., 2021). This argument is contradicted by the results of a study by Karayannis et al. (2013) who found that fear of movement assessed with the TSK was linked to rigidity of the trunk, but not an item that was designed to capture fear of the task used for the rigidity assessment. Nevertheless, the use of more specific measures of fear may be relevant to the clarification of the relationship between fear of movement and postural sway. Therefore, we apply this concept of more specific fear of movement assessments to study the relation with postural sway.

Separate variables are often used to describe sway in the sagittal and the frontal plane, as for example in the study of Mazaheri et al. (2014). This distinction is relevant, because the mechanisms central for regulating postural sway in different planes partially differ (Winter, 1995). In contrast, fear of movement is usually assessed in a more general format and no distinction between movement planes is made. Although the Photograph Series of Daily Activities-Short Electronic Version (PHODA-SEV) was designed to comprise of movement examples for different planes, the resulting score does not distinguish between these planes (Leeuw et al., 2007). As seen in the study of Leeuw et al. (2007) not all movements are perceived as equally harmful. Therefore, on a slightly more general level, for some people fear of bending the trunk in the sagittal plane may differ from fear of bending the trunk in the frontal plane. Fear of bending the trunk in different planes could have distinct effects on postural sway. Direction specific fears might result, for example, in a restriction of sway in the movement plane corresponding to the fear. We assumed that fears of bending the trunk in different planes might relate differently to postural sway in different planes.

To gain further insights into the relationship between fear of movement and postural sway we conducted secondary analyses of COP data. This data was obtained from people with LBP who participated in a clinical trial that investigated the effect of an exercise intervention (Meinke et al., 2022). Our aim was to investigate whether postural sway, described by mean displacement and velocity, is related to fear of movement in general, and whether fear of bending the trunk in different planes relate to sway for the corresponding movement directions.

## 2 Materials and methods

### 2.1 Study design and participants

We describe secondary analyses of data from a randomized controlled trial which investigated the effects of an exergame for people with LBP on postural sway (Meinke et al., 2021, 2022). Data of 2 baseline assessments taken about 3 weeks apart were used (visit 1 and 2). The first visit was completed by 32 participants and 27 participants completed the second visit. A total of 59 observations of postural sway and general fear of movement were available. As the directional questions had been introduced after an amendment to the study protocol, the data used for directional analyses originates from 25 unique participants. Of these 25 participants, 16 contributed data for both visits. Participants had not yet been randomized or received any intervention at the time the data were captured. The prospective study participants were contacted using leaflets, online advertisement, and personal interaction. Participants were included if they had back pain in the lower region, were above 18 years old, not in any therapies for LBP, for the past half year and gave their informed consent. Participants were excluded from the study due to radicular symptoms or other specific causes of LBP, when vision was too low to use the exergame, when the participants were in pharmacological treatment that negatively influences sway or when they had allergies to the band used to adhere sensors to the skin. We further excluded pregnant women, or participants for whom language barriers did not permit participation. In addition, we informed the participants about the nature of the exercise program and asked to self-assess whether their pain would allow to perform these movements. This was asked to exclude participants who could have not completed the exercises due to strong pain. The study was conducted according to the current Declaration of Helsinki and ethical approval was received from the Cantonal Ethics Committee Zurich, Switzerland (BASEC-2018-02132). Further information on procedures and materials not directly relevant for addressing the research question at hand is available in (Meinke et al., 2021, 2022).

### 2.2 Procedures

Fear of movement, postural sway and pain intensity data were collected at both assessment visits. A numeric rating scale (0-10) was used to query pain intensity during the past 7 days (Chiarotto et al., 2018). Weight and height were determined at the first assessment. Fear of movement was quantified among other variables using an online form through RedCap (Harris et al., 2009). All participants took the survey on the same computer monitor before the postural sway assessments were carried out. The questions were either presented in German or English.

### 2.3 Fear of movement

For a generic score of fear of movement, we used an abbreviated version of the Tampa scale for Kinesiophobia (TSK-11) (Woby et al., 2005; Rusu et al., 2014). The English assessment shows good psychometric quality, which is comparable to the full scale in patients with chronic LBP (Woby et al., 2005). The German TSK-11 has been validated and shows sufficient internal consistency (Rusu et al., 2014). Direction specific items for fear of movement (in the following referred to as *directional fear*) were included later during the study, therefore data of fewer participants were available. For this directional fear assessment, a custom question type was used. Earlier studies, which had integrated fear assessments more directly related to the movements under investigation, appended further items to the PHODA-SEV (Karayannis et al., 2013) or used items already available within the PHODA-SEV (Matheve et al., 2019; Knechtle et al., 2021). Similar to the style of the PHODA-SEV, the participants rated how harmful they perceive bending the trunk in different directions (Figure 1). As the PHODA-SEV requires the participants to put photographs in order depicting different movements, we added icons to clarify the movement referenced in the questions. Fear of bending the trunk in the sagittal plane was defined as the mean of two items referring to trunk flexion and extension in the sagittal plane. A measure to quantify whether the participants judged fear of bending the trunk in the frontal plane or sagittal plane trunk as higher was introduced and is hereinafter referred to as *relative directional fear*. Relative directional fear was calculated by subtracting fear of bending the trunk in the frontal plane from fear of bending the trunk in the sagittal plane. Negative values describe stronger fear of bending the trunk in the frontal plane, while positive values describe relatively stronger fears of bending the trunk in the sagittal plane. Values close to zero indicate no difference in fear ratings between frontal and sagittal plane movements. For example, if a participant has low values in the sagittal and the frontal plane, this results in a relative directional fear value of zero. On the other hand, if a participant has high values in the sagittal and the frontal plane, this results in a relative directional fear value of zero as well. This assessment is therefore a measure of similarity of the plane ratings and the direction in which fear is *relatively* higher, rather than a measure for the general level of fear of bending the trunk.

**Figure 1:**
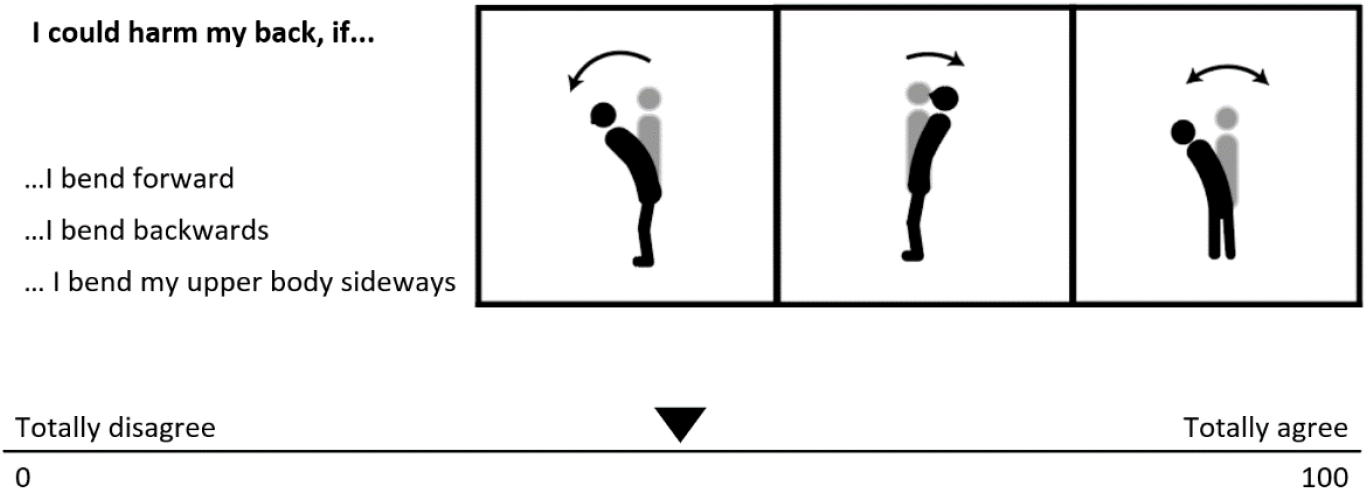
Question format used for the assessment of directional fear.

### 2.4 Assessment of postural sway

The assessments of postural sway were implemented considering the suggestions from Ruhe et al. (2010) and were performed for 120 seconds in 4 consecutive repetitions. COP data was acquired from a pressure plate (AMTI, Accusway Plus, Watertown, MA, USA). The participants maintained still, upright stance and had their hands loosely hanging, closed their eyes during the assessments, and wore shaded ski goggles. Measurements were performed removing visual information while standing on a stable surface because this had been recommended to improve the reliability (Ruhe et al., 2010). The exact stance position of the participants was outlined on a synthetic sheet to replicate the stance throughout all subsequent recordings. Prior to and after each repeat, participants performed a movement leaning to both sides. This was necessary for combining the recordings with data from other sensors that were not relevant for the analyses presented here. The movement was removed including the 5 seconds after and before the movement respectively. Thus, of each trial only 110 seconds were analyzed. After filtering the COP recordings with a low-pass fourth order Butterworth filter (10 Hz cut-off frequency) and centering the data, displacement from the center, velocity and their directional versions were calculated according to Prieto et al. (1996) (equation 1-5, 8-11). General displacement was calculated by averaging the distances of each data point from the center of the trajectory. In an analogous way, displacement for the sagittal and frontal plane were calculated by averaging the absolute values of the x or y coordinates of each point in the trajectory. General velocity was calculated as the path length traveled divided by time, and velocities for the sagittal and frontal plane were calculated by the path traveled on the respective axis divided by time. These preparatory steps were performed using MATLAB R2018a (The MathWorks Inc., Natick, MA, USA). Averages across the 4 trials were used for data analysis.

### 2.5 Statistical methods

For hypothesis testing, we used mixed effects linear models. R (R Core Team, 2021) version 4.04 and the package *lme4* (Bates et al., 2015) were used for the statistical analysis. Continuous predictors including pain scores and fear of movement assessments were standardized across the entire sample before inclusion into the models. Normal distribution of the residuals and random intercepts were assessed using normal-qq plots and the presence of heteroscedasticity was reviewed by plotting the predicted values against the residuals. The residuals were further plotted against each predictor variable individually. These analyses were performed for the untransformed, log(x+1), square root transformed and reciprocally transformed data. Based on the residual analysis the models with log transformed outcomes were chosen. For each postural sway outcome, we estimated a first model to assess the contribution of potential covariates (assessment visit, sex, age, height, weight and pain). Based on the results of this analysis of covariates, we included the variables visit, age and weight as fixed effects in the baseline models for all sway outcomes. The associations of the fear of movement variables with the postural sway variables were tested by adding each individual variable of interest to the baseline model and comparing the resulting model against the baseline model only. To compare the models, we performed Likelihood-ratio tests by using the ANOVA function as described in Bates et al. (2015). All models included the participants as random effect. We used α=.05 as a threshold for accepting statistical significance. The influence of every participant on the models was assessed using Cook’s distance calculated using the R package *Influence*.*ME* (Nieuwenhuis et al., 2012). As suggested in Nieuwenhuis et al. (2012), a threshold of 4 divided by the number of participants in the model was used. All variables of the respective model were included. Each participant classified as influential in the initial model was separately removed from the models and comparisons were reevaluated to see if individual participants affected the statistical significance of the results. Spearman correlation coefficients for fear of movement variables and directional postural sway variables were calculated. Reliability of fear of movement assessments and postural sway assessments was estimated using ICC model (2,1), relying on the ANOVA results as implemented in the package *psych*, version 2.2.3 (Revelle, 2021) and interpreted according to the reliability thresholds proposed by Portney and Watkins (1993). Reliability estimates were based on data from participants who completed the questionnaires at both assessment visits. For one participant the values for directional fear were missing and thus imputed with the value of 50, representing the default slider position. Replacements were made as we assumed an agreement with the default slider position (not producing an output value if the slider was not clicked) was more plausible, than the participant having overlooked these questions.

## 3 Results

### 3.1 Participant characteristics

Directional fear was obtained from 20 observations in visit 1 and 21 observations in visit 2 (Table 1). Participant characteristics, general and directional fear of movement, and postural sway estimates from both visits are summarized in Table 1. In many cases, ratings of fear of bending the trunk in the frontal and in the sagittal plane were similar (Figure 2A, values close to the diagonal line). Participants reporting relatively higher fear of bending the trunk in the sagittal plane showed rather low values for both directions (Figure 2A, dashed triangle). Relative directional fear was calculated by subtraction of fear of bending the trunk in the frontal plane from fear of bending the trunk in the sagittal plane, resulting in a distribution centered around zero (Figure 2B). Negative values indicated higher fear of bending the trunk in the frontal plane whereas positive values showed that fear of bending the trunk was rated higher for the sagittal plane. Estimates of intra-class-correlation (ICC) coefficients for directional fear variables were in the range from moderate to good (Table 2). However, several of the confidence intervals included values below 0.5 and in these cases we could not exclude poor reliability (Table 2).

**Table 1.** Descriptive statistics of participant characteristics, fear assessments and postural sway outcomes at both assessment visits.

| Variable | Visit 1 | Visit 2 |
| --- | --- | --- |
|  | Median (IQR)<sup>a</sup>/ M (SD)<sup>b</sup> | Median (IQR)/ M (SD) |
| Participant characteristics |  |  |
| N | 32<sup>c</sup> | 27<sup>c</sup> |
| Age | 37.50 (24.75)<sup>a</sup> | 35.00 (25.5)<sup>a</sup> |
| Height | 171.26 (7.92)<sup>b</sup> | 172.05 (7.65)<sup>b</sup> |
| Sex (male/female) | 11/21<sup>c</sup> | 10/17<sup>c</sup> |
| Language (English/German) | 6/26<sup>c</sup> | 5/22<sup>c</sup> |
| Pain Intensity | 3.19 (1.47)<sup>b</sup> | 2.59 (1.34)<sup>b</sup> |
| General Fear |  |  |
| TSK-11 | 19.59 (4.41)<sup>b</sup> | 19.96 (5.62)<sup>b</sup> |
| Displacement (mm) | 4.82 (1.71)<sup>a</sup> | 5.24 (2.13)<sup>a</sup> |
| Velocity (mm/s) | 9.76 (4.11)<sup>a</sup> | 11.15 (4.42)<sup>a</sup> |
| Directional Fear |  |  |
| n | 20<sup>c</sup> | 21<sup>c</sup> |
| Fear flexion | 23.00 (50)<sup>a</sup> | 18.00 (26)<sup>a</sup> |
| Fear extension | 31.50 (49.5)<sup>a</sup> | 17.00 (49)<sup>a</sup> |
| Fear sagittal | 32.00 (46.75)<sup>a</sup> | 18.50 (39.5)<sup>a</sup> |
| Fear frontal | 25.00 (47.5)<sup>a</sup> | 17.00 (25)<sup>a</sup> |
| Relative directional fear | 0.00 (6.25)<sup>a</sup> | 0.00 (17.5)<sup>a</sup> |
| Postural Sway |  |  |
| Displacement sagittal (mm) | 4.14 (1.71)<sup>a</sup> | 4.22 (1.90)<sup>a</sup> |
| Displacement frontal (mm) | 1.88 (1.53)<sup>a</sup> | 2.54 (1.35)<sup>a</sup> |
| Velocity sagittal (mm/s) | 7.86 (4.24)<sup>a</sup> | 9.36 (4.04)<sup>a</sup> |
| Velocity frontal (mm/s) | 4.48 (1.95)<sup>a</sup> | 3.77 (2.60)<sup>a</sup> |

<sup>a</sup> Median (IQR): Median (inter quartile range).
<sup>b</sup> M (SD): Mean (standard deviation).
<sup>c</sup> Count: Counted data

**Table 2.** Reliability estimates for directional fear questions (*n*=16) and postural sway measures (*n*=27).

| Variable | Intraclass Correlation Coefficient |
| --- | --- |
|  | Estimate (95% CI) |
| Fear flexion | 0.79 (0.50 to 0.92) |
| Fear extension | 0.59 (0.16 to 0.83) |
| Fear sagittal | 0.68 (0.30 to 0.88) |
| Fear frontal | 0.74 (0.40 to 0.90) |
| Relative directional fear | 0.84 (0.61 to 0.94) |
| Displacement | 0.77 (0.55 to 0.89) |
| Displacement sagittal | 0.77 (0.56 to 0.89) |
| Displacement frontal | 0.64 (0.35 to 0.82) |
| Velocity | 0.77 (0.56 to 0.89) |
| Velocity sagittal | 0.78 (0.57 to 0.89) |
| Velocity frontal | 0.79 (0.59 to 0.90) |

**Figure 2:**
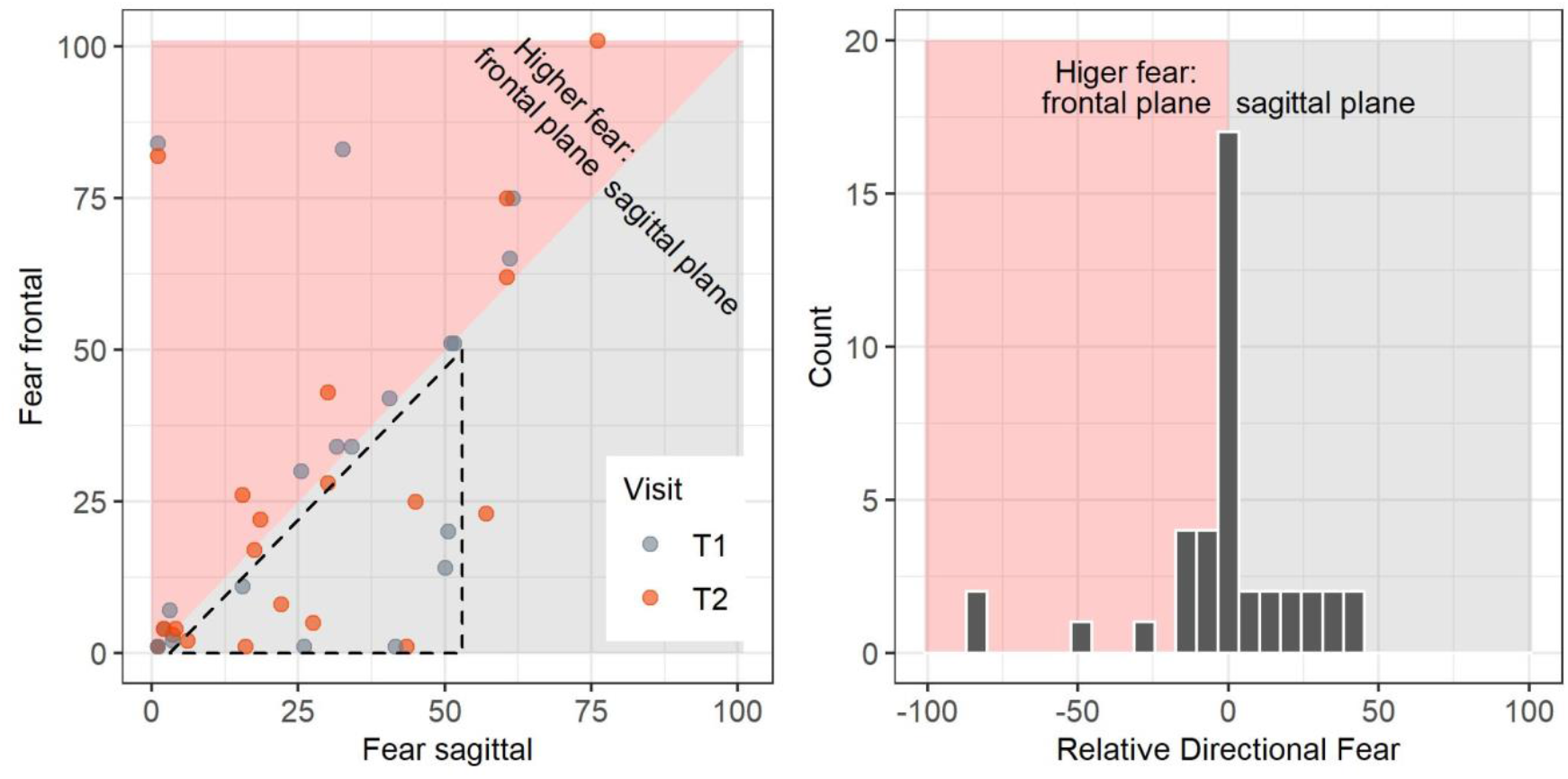
Relative directional fear is calculated by subtracting fear ratings for the frontal plane from fear ratings of the sagittal plane. Background in red shade indicates higher fear values for the frontal plane, grey shade background shows area of higher fear in the sagittal plane. **(A)** Ratings of fear in the sagittal and the frontal plane. The dashed triangle highlights that participants who rated fear in the sagittal plane higher than fear in the frontal plane showed low to medium values on both planes. **(B)** Distribution of relative directional fear. Negative values for relative directional fear show higher fear of frontal plane movements and positive values show higher fear of sagittal plane movements.

### 3.2 Association of fear of movement with postural sway

The assessment of covariates in baseline models resulted in assessment occasion, age and weight to be included in the baseline models for all outcomes (Table 3). Comparisons of the baseline models including the fear predictor of interest to the baseline model alone are reported in Table 4. General fear of movement as measured by the TSK-11 was not associated with general mean displacement and velocity. However, for the directional sway measures, an association was found with velocity of sway in the frontal plane. Fear of bending the trunk in the sagittal plane was not associated with any of the directional sway measures. Fear of bending the trunk in the frontal plane was associated with displacement and velocity in the frontal plane, but also with displacement in the sagittal plane. The predictor relative directional fear showed the same pattern of associations.

**Table 3.** Models assessing the contribution of covariates and baseline models only including the covariates selected as relevant.

| Predictor | Outcomes<br>Displacement | Velocity | Displacement<br>Sagittal | Frontal | Velocity<br>Sagittal | Frontal |
| --- | --- | --- | --- | --- | --- | --- |
| *n* comparison | 32 | 32 | 25 | 25 | 25 | 25 |
| **Covariates** |  |  |  |  |  |  |
| Visit | 0.07 (-0.00 to 0.13) | 0.04 (-0.02 to 0.10) | 0.03 (-0.07 to 0.11) | 0.16 (0.05 to 0.27) | 0.07 (-0.01 to 0.15) | 0.06 (-0.02 to 0.14) |
| Sex | 0.04 (-0.19 to 0.27) | -0.02 (-0.21 to 0.17) | 0.07 (-0.13 to 0.26) | 0.14 (-0.14 to 0.43) | 0.02 (-0.19 to 0.22) | 0.05 (-0.18 to 0.29) |
| Age | 0.01 (-0.08 to 0.10) | 0.08 (0.01 to 0.16) | -0.05 (-0.13 to 0.03) | 0.09 (-0.03 to 0.21) | 0.09 (0.01 to 0.17) | 0.04 (-0.06 to 0.13) |
| Height | 0.04 (-0.08 to 0.15) | 0.03 (-0.07 to 0.12) | -0.07 (-0.18 to 0.03) | -0.02 (-0.17 to 0.13) | -0.02 (-0.12 to 0.09) | -0.03 (-0.15 to 0.09) |
| Weight | -0.05 (-0.14 to 0.04) | 0.05 (-0.02 to 0.13) | 0.04 (-0.05 to 0.13) | -0.01 (-0.14 to 0.12) | 0.09 (-0.00 to 0.19) | 0.08 (-0.02 to 0.19) |
| Pain Intensity | 0.02 (-0.03 to 0.07) | 0.01 (-0.03 to 0.06) | -0.00 (-0.06 to 0.05) | 0.04 (-0.03 to 0.11) | 0.01 (-0.05 to 0.06) | 0.03 (-0.03 to 0.08) |
| **Baseline Model** |  |  |  |  |  |  |
| Visit | 0.06 (-0.01 to 0.12) | 0.03 (-0.02 to 0.09) | 0.03 (-0.06 to 0.11) | 0.13 (0.03 to 0.24) | 0.07 (-0.01 to 0.14) | 0.04 (-0.04 to 0.12) |
| Age | 0.01 (-0.08 to 0.10) | 0.08 (0.00 to 0.15) | -0.04 (-0.13 to 0.04) | 0.10 (-0.02 to 0.22) | 0.09 (0.01 to 0.18) | 0.05 (-0.05 to 0.14) |
| Weight | -0.04 (-0.13 to 0.05) | 0.06 (-0.01 to 0.13) | 0.01 (-0.09 to 0.10) | -0.03(-0.16 to 0.11) | 0.09 (-0.00 to 0.18) | 0.07 (-0.03 to 0.18) |

Estimate and (95% CI).
All estimates are based on log transformed outcome variables. Continuous predictors (including pain intensity) were standardized before analysis. Statistically significant results are in bold type.

**Table 4.**
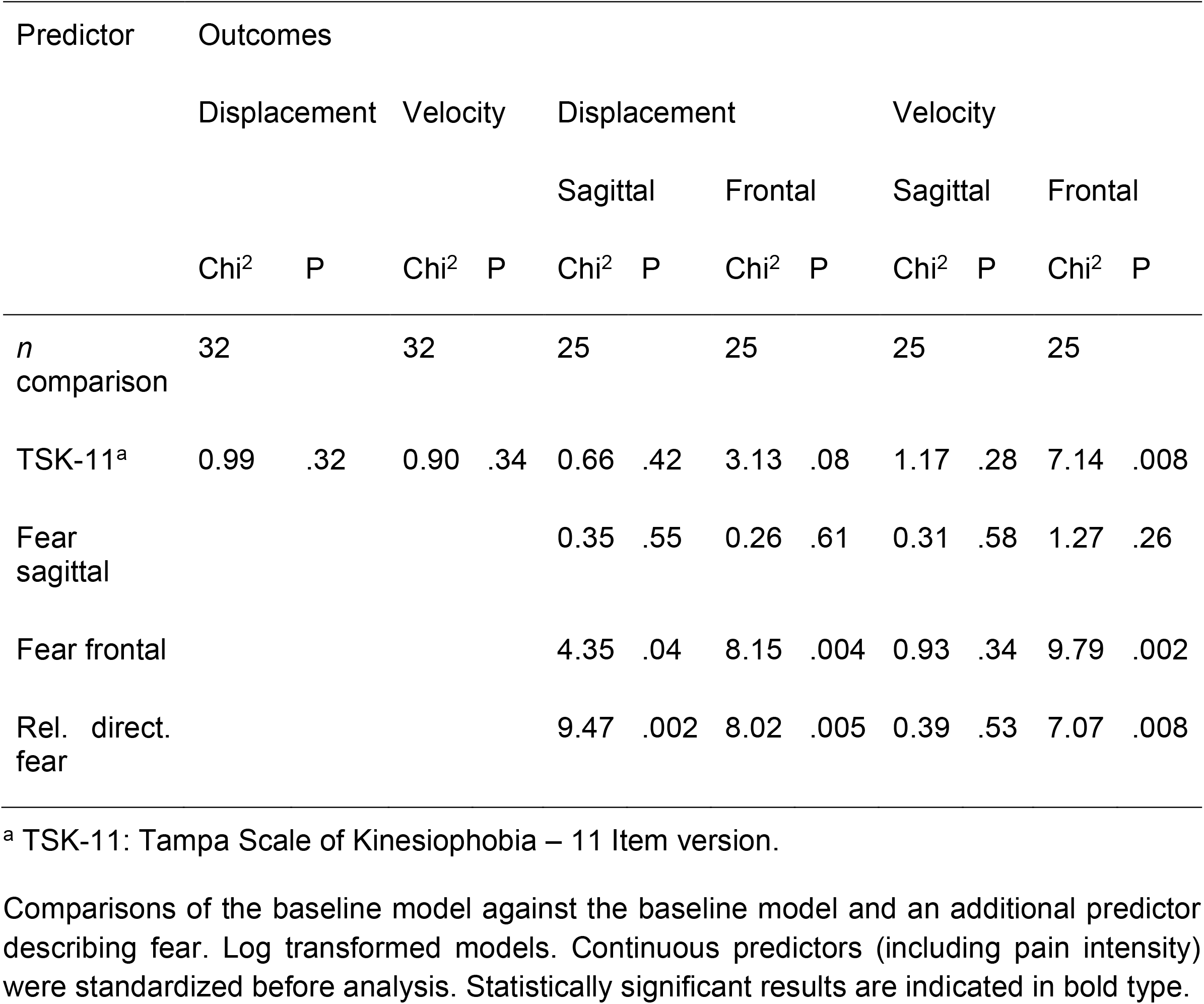
Comparisons between the baseline model and the baseline model including an additional fear predictor variable.

We tested whether removing individual participants that had been identified as influential by using Cook’s distance would have changed the results of the model comparisons. For two cases, when either of these participants would be removed, the association of fear of bending the trunk in the frontal plane with displacement in the sagittal plane was no longer significant (both with *p*=.05). In addition, the association of general fear of movement with displacement in the frontal plane could have become statistically significant by removing one influential case.

Graphical representations (Figures 3A-H and 4A-H) of the untransformed directional data showed that displacement and velocity both tended to be rather higher than lower with higher fear of movements. The graphical displays for relative directional fear showed that participants with *relatively* higher fear of bending the trunk in the frontal plane tended toward higher displacement in both directions, whereas participants with *relatively* higher fear of bending the trunk in the sagittal plane had average to low displacements. Spearman correlation coefficients describing the associations of fear variables with directional sway measures at both assessments are reported in supplementary file 1.

**Figure 3:**
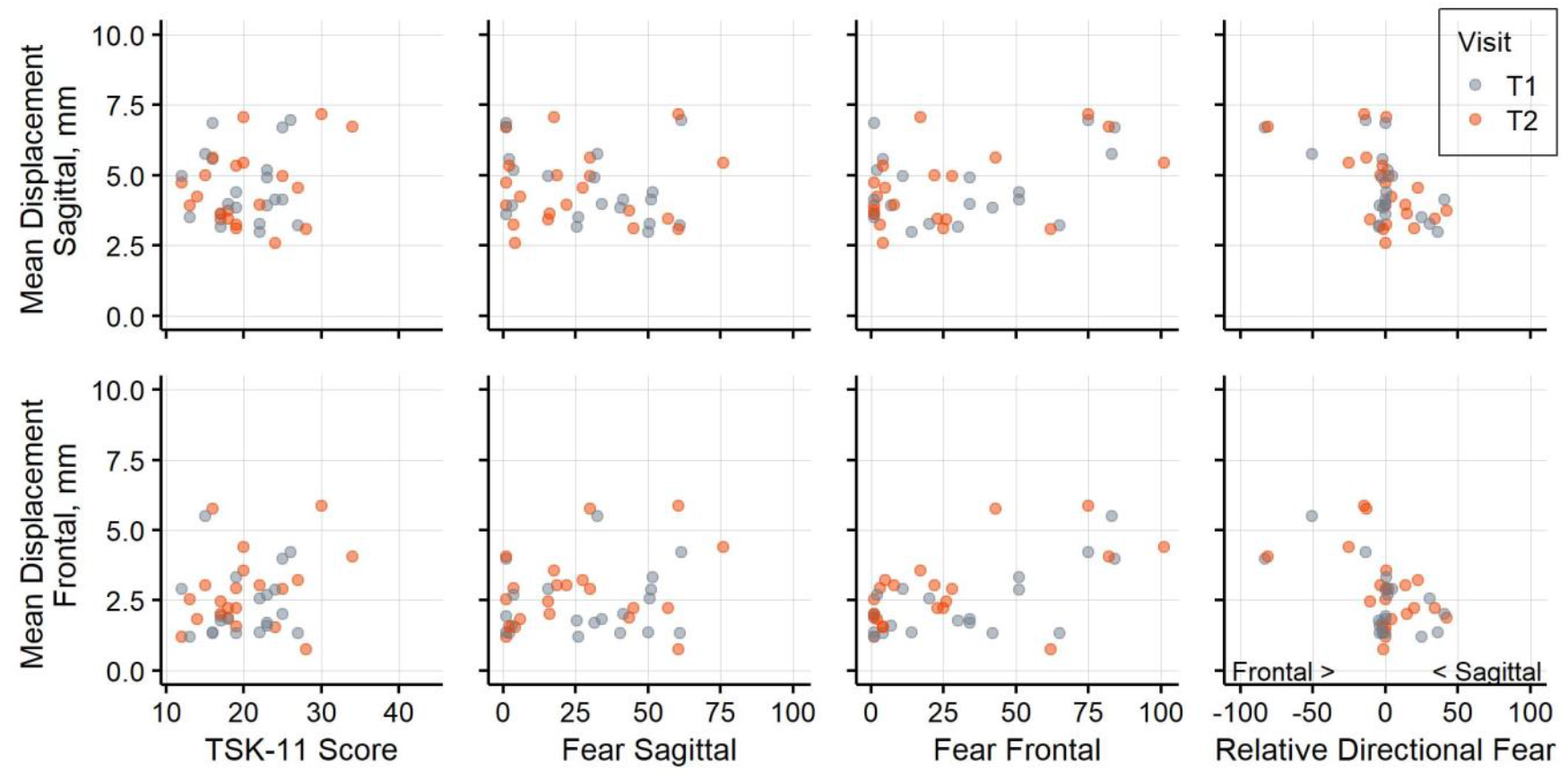
Postural sway displacement and fear variables. Association of postural sway displacement in the sagittal plane and **(A)** general fear, **(B)** fear of sagittal plane movement, **(C)** fear of frontal plane movement, **(D)** relative directional fear. Association of postural sway displacement in the frontal plane and **€** general fear, **(F)** fear of sagittal plane movement, **(G)** fear of frontal plane movement, **(H)** relative directional fear. Negative values for relative directional fear show higher fear of frontal plane movements and positive values show higher fear of sagittal plane movements. Data from assessment visit T1 is shown in blue and data from T2 in orange.

Graphical inspection revealed that of the movements comprising the measure for fear of bending the trunk in the sagittal plane, flexion and extension, flexion considered separately appear to have a positive association with velocity measures (Supplementary file 2). Therefore, additional model comparisons for flexion and extension were conducted. Neither flexion nor extension in the sagittal plane had a statistically significant association with any of the directional postural sway outcomes.

## 4 Discussion

### 4.1 Summary of main findings

We conduced analyses on the association of fear of movement and postural sway considering fear of movement in general and fear of bending the trunk in different planes. The TSK-11, a measure of general fear of movement, was not associated with undirected measures of sway, but was related to velocity in the frontal plane. Other than expected, fear of bending the trunk in individual planes were not more closely associated with sway measures for the corresponding plane. Instead, fear of bending the trunk in the sagittal plane was not associated with any directional outcome. Fear of bending the trunk in the frontal plane was associated with displacement in the frontal and the sagittal plane and with velocity in the frontal plane. Sway parameters tended to increase rather than decrease with elevated fear.

### 4.2 Association of fear of movement and postural sway

It had been proposed that assessments of fear of movement should be based on concrete movement examples rather than general assessments (Leeuw et al., 2007) and that assessments of fear of movement which match more precisely with the movement task, might be better suited to detect associations with movement quality (Pincus et al., 2010; Matheve et al., 2019). Our results support this notion only partially, as we observed a significant association between general fear of movement and sway velocity in the frontal plane. As we detected more associations of directional fear questions with sway outcomes, the question format encouraging participants to think of specific movements could have contributed to making an association between fear of movement and sway more visible.

The results of the directional analysis showed that fear of bending the trunk in the frontal plane, but not fear of bending the trunk in the sagittal plane, was associated with postural sway in people with LBP. A potential explanation of these results may be derived from the basic mechanisms at work during bipedal stance. During regular standing with approximately parallel feet on stable ground, two basic mechanisms have been suggested as central to regulating postural sway (Winter, 1995). Balance control in the frontal plane should rely on muscle contractions at the level of the pelvis, moving the body weight laterally, whereas sway in the sagittal plane is regulated predominantly by contractions at the level of the ankle (Winter, 1995). Fear of bending the trunk in the frontal plane might thus interfere with the use of this lateral weight shifting of the hip. By contrast, less movement of the trunk or the hip in the sagittal plane should be required for regulating balance under these basic stance conditions. Therefore, fear of bending the trunk in the sagittal plane might cause less interference with balance under the investigated conditions. The results of Mok et al. (2004) suggest that people with LBP could rely less on their hip for regulating sway. Furthermore, Mok et al. (2004) noticed that fear of movement might be a factor related to limiting the use of the hip. Nonetheless, a reduction in velocity and a descriptive tendency towards increased range in the sagittal plane in some conditions was observed in people with LBP (Mok et al., 2004). To note, our work did not focus on testing the assumption that the observed pattern of results might be caused by mechanisms involving the hip and future studies are required.

### 4.3 Direction of the association between fear of movement and postural sway

The data obtained in this study suggests a positive association, where elevated fear of movement lead to an increase in sway parameters rather than a decrease. It could be argued that people with LBP and high fear of movement might refrain from using an effective mechanism for balance regulation (i.e., the lateral weight shifting described above) which could compromise balance and lead to an increase in sway. This argumentation is comparable to the view presented in the review of Koch and Hänsel (2019). They stated that postural sway is increased under more challenging conditions in people with LBP, potentially because the hip strategy required for managing these conditions could be impaired. In contrast, others have argued that elevated pain-related fear might be linked to a decrease of sway parameters (Mazaheri et al., 2013, 2014; Kiers et al., 2015). This assumption of decreased sway is supported by empirical data from a study of Shanbehzadeh et al. (2018), which suggests less sway in people with elevated pain-related fear in comparison with people with lower fear. Methodological factors that differ from the approach used in our study include the categorization of participants with low and high fear based on the Pain Anxiety Symptom Scale-20, the standing condition with the feet positioned directly next to each other and the use of multiple assessment conditions including for example dual task assessments (Shanbehzadeh et al., 2018). These aspects may have played a role causing the differences between the observed results. Other researchers who assessed the association of fear of movement or similar concepts and postural sway under bipedal stance conditions without platform perturbations obtained deviating results (Mazaheri et al., 2014; Kahraman et al., 2018; Zhang et al., 2020; Mikkonen et al., 2022). Mazaheri et al. (2014) concluded that fear of movement did have little effect but did not report the association of sway and fear directly. Instead, to infer the effect of fear of movement, based on the differences in pain and fear of movement status they relied on comparisons between groups of people with LBP, people who just recovered, and healthy participants (Mazaheri et al., 2014). Mikkonen et al. (2022) arrived at the same conclusion, that fear of movement was not associated with postural sway, but nevertheless had reported several positive correlations of fear of movement with sway area measures. Kahraman et al. (2018) found an increase in postural sway only for male participants and only for a score combining assessments from several conditions, but otherwise found no significant correlations. Zhang et al. (2020) supposedly found larger sway for participants with higher catastrophic thoughts, but the direction of the detected association was not reported consistently throughout the manuscript. Thus, our study complements this existing literature with an estimate for a tendency towards increased sway with higher fear. The opposing findings in the currently available literature could be caused by the different fear concepts and their operationalizations, as well as variations in balance assessments.

### 4.4 Relative directional fear and postural sway

Values for relative directional fear were closely centered around zero and many participants did not judge one movement direction as more harmful than the other. This suggests that the movement plane with respect to fear may be only relevant for a smaller number of people. We found several statistically significant associations between relative directional fear and directional sway variables in the model comparisons. As presented in Figure 3, relatively higher fear of bending the trunk in the frontal plane was associated with higher displacements, while relatively higher fear of bending the trunk in the sagittal plane corresponded to average or low displacements. However, the results on relative directional fear need to be interpreted with caution. Figure 2 A shows that participants with higher fear of bending the trunk in the sagittal plane tended to have lower values for both planes, whereas participants with higher fear of bending the trunk in the frontal plane often rated both planes higher. This pattern indicates that in this dataset, relative directional fear was conflated to some degree with the general tendency to rate fear as higher or lower. Therefore, these analyses should be repeated based on other datasets before conclusions on relative directional fear should be drawn.

### 4.5 Strengths and Limitations

Several methodological aspects must be considered when interpreting the current results. The data originated from study participants with LBP who registered for a trial investigating an exercise intervention. Therefore, a selection bias might be present. For example, only participants with lower levels of fear of movement may have registered for the trial and the results need to be interpreted in the context of this subset. Furthermore, to assess the directional fear, we introduced custom questions that did not originate from a validated questionnaire. The ICC estimates calculated for these questions indicated moderate to good reliability. Nevertheless, these estimates were derived from a smaller number of participants and thus had broader confidence intervals which could partially not rule out poor reliability. In addition, it remains unclear whether it was adequate to use the mean of the flexion and extension question as a measure for fear of bending the trunk in the sagittal plane. In a recent fear learning experiment, fear of extension of the trunk was rated higher than fear of flexion, but the images showed a more extreme extension movement than it was depicted in this study (Gatzounis et al., 2021). Visual inspection of our data indicated that there may be an association of postural sway velocity with flexion movements, but not extension (Supplementary File 2). Future studies may therefore need to consider flexion and extension in the sagittal plane separately. However, no statistically significant association of fear for flexion or extension in the sagittal plane with directional postural sway measures were found when the corresponding model comparisons controlling for age and weight were performed in addition. As described above, the results were mostly robust to the deletion of individual participants from the analyses. Nevertheless, Figure 3 (A-H) and Figure 4 (A-H) show that the results seem to be determined largely by a small number of participants, and that many participants reported no or very little fear of bending the trunk in both planes. Furthermore, many statistical tests were performed in this analysis, and we did not adapt the significance thresholds to counteract an inflation of the error probability. In addition, the analyses were designed to detect an association between fear of movement and sway variables. They do not exclude the possibility that an association was present in cases where we could not detect an association.

**Figure 4:**
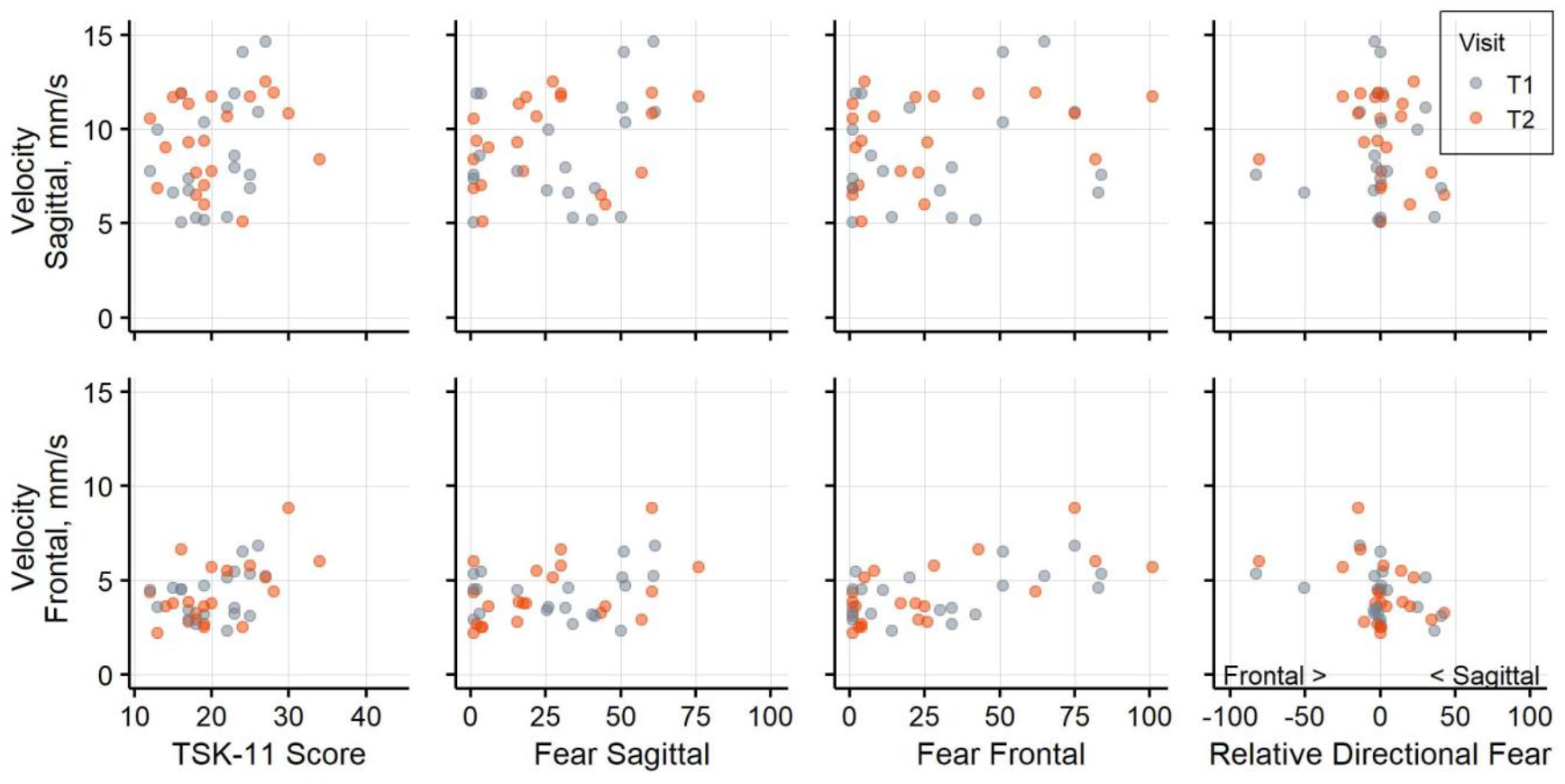
Postural sway velocity and fear. Association of postural sway velocity in the sagittal plane and **(A)** general fear, **(B)** fear of sagittal plane movement, **(C)** fear of frontal plane movement, **(D)** relative directional fear. Association of postural sway velocity in the frontal plane a€**(E)** general fear, **(F)** fear of sagittal plane movement, **(G)** fear of frontal plane movement, **(H)** relative directional fear. Negative values for relative directional fear show higher fear of frontal plane movements and positive values show higher fear of sagittal plane movements. Data from assessment visit T1 is shown in blue and data from T2 in orange.

Other methodological considerations concern the variables that could be controlled in this study. The analyses were complex and more robust estimates should be obtained from larger datasets in future studies. For instance, without including age and weight in the models, the association of fear of bending in the frontal plane with displacement in the sagittal plane would not have been statistically significant. In the corresponding baseline model, the association with age was estimated to be negative (Table 3), although a positive association with age would be expected and was found for other sway variables. Furthermore, when flexion and extension ratings were considered separately, the association of flexion with velocity became significant in both planes. However, the association of age with postural sway is well established (Roman-Liu, 2018), and therefore controlling for the influence of age is important. In addition to fear of movement, pain and how careful movements would be performed were assessed using directional type questions as well. These data were collected, as it is important to discern how movements are avoided and how painful they are experienced (Pincus et al., 2010). Unfortunately, the available number of participants did not permit to include these predictors and we aimed to maintain the comparability of the baseline models between the different outcomes. Although it has been reported that pain did not account for the link between pain-related fears and rather stiff movement of the spine (Christe et al., 2021), future studies should include pain in a directional format as well and not only consider pain on a general level. The directional fear of movement assessment queries the perceived harmfulness of bending the trunk in the frontal or sagittal plane using individual items. Naturally, these items cover the construct *fear of movement* in less depth than the more complex TSK. Therefore, the directional assessment can to some degree differ conceptually from the general fear assessment, other than with respect to the directional nature. Additionally, even though we consider the standing task in this study as easy for most participants, in future studies fear of falling during the task should be assessed. This would facilitate comparisons between studies using different postural sway assessments.

### 4.6 Conclusion

To our knowledge this is the first study exploring the associations of directional fear of movement with postural sway in people with LBP. The presented data suggest that fear of bending the trunk in the frontal plane may be positively associated with several measures of postural sway in people with LBP under the investigated stance conditions. We hope these preliminary results can draw further attention to the need to match the level of abstraction of the fear assessment to the level of the movement analysis. Continued work should replicate these results, validate the format of the questions used, and pursue exploration of the mechanisms underlying these observations.

## Data Availability

Raw data that support the findings of this study cannot be made publicly available to protect participants' rights according to Swiss human research law. The deidentified individual participant data that underly the results of this paper can be
accessed by investigators who (1) submit a methodological sound proposal describing the intended analysis and as reviewed by the authors of this publication, (2) provide proof of relevant ethical approval for the intended analysis, and (3) fulfill data protection measures according to Swiss legal requirements.

## 4.7 Acknowledgements

We would like to thank all the participants and the people who supported this study. Tina Wunderlin, Ramon Glättli, Katharina Zahoranszky, Kim Graf and Adrian Stutz helped with performing postural sway assessments with the participants. Ruud Knols and Rick Peters were involved in conducting the primary randomized controlled trial and in the examination of eligibility of study participants. Laura Tüshaus reviewed the custom fear questions. Joanne Lim provided feedback on the manuscript. We received advice regarding the statistical analyses from the Seminar for Statistics at ETH Zurich.

## 6 Author Contributions

AM, CM, MM, JS and WK planned this work. CM participated in the data collection. AM managed the study, analysed the data, and prepared the first version of the manuscript. AM, MM, JS and WK edited the manuscript. All authors consent to the publication of the manuscript in its current form.

